# A Targeted Approach for Evaluating DUX4-Regulated Proteins as Potential Serum Biomarkers for Facioscapulohumeral Muscular Dystrophy Using Immunoassay Proteomics

**DOI:** 10.1101/2022.07.25.22276982

**Authors:** Amy E. Campbell, Jamshid Arjomand, Oliver D. King, Rabi Tawil, Sujatha Jagannathan

## Abstract

**Background:** Facioscapulohumeral muscular dystrophy (FSHD) is a progressive myopathy caused by misexpression of the double homeobox 4 (DUX4) embryonic transcription factor in skeletal muscle. Identifying quantitative and minimally invasive FSHD biomarkers to report on DUX4 activity will significantly accelerate therapeutic development.

**Objective:** The goal of this study was to analyze secreted proteins already known to be induced by DUX4 in order to identify potential blood-based molecular FSHD biomarkers.

**Methods:** We used high-throughput, multiplex immunoassays from Olink Proteomics to measure the levels of several known DUX4-induced genes in a cellular myoblast model of FSHD, in FSHD patient-derived myotube cell cultures, and in serum from individuals with FSHD. Levels of other proteins on the Olink Proteomics panels containing these DUX4 targets were also examined in secondary exploratory analysis.

**Results:** Placental alkaline phosphatase (ALPP) levels correlated with DUX4 expression in both cell-based FSHD systems but did not distinguish FSHD patient serum from healthy controls.

**Conclusions:** ALPP, as measured with the Olink Proteomics platform, is not a promising FSHD serum biomarker candidate but could be utilized to evaluate DUX4 activity in discovery research efforts.

## INTRODUCTION

Facioscapulohumeral muscular dystrophy (FSHD) is an inherited myopathy with no disease-modifying or curative treatment options [1]. FSHD is caused by misexpression of the *double homeobox 4* (*DUX4*) gene in skeletal muscle following epigenetic de-repression of the D4Z4 macrosatellite repeat array on chromosome 4q35 due to D4Z4 array contraction (type 1) or mutations in genes encoding D4Z4 chromatin modifiers (type 2) [2–5]. Sustained misexpression of DUX4, a transcription factor important for early embryonic development, is toxic [6–9]. Toxicity is thought to be caused by the dysregulation of genes and proteins involved in germline and stem cell development, myogenesis, innate immunity, and several other important cellular functions, though exactly how downstream molecular changes lead to pathology is not fully understood [10, 11]. In individuals with FSHD, DUX4-induced cell death leads to skeletal muscle fiber degeneration and replacement with fatty, fibrous tissue resulting in progressive muscle weakness and disabling physical limitations. Skeletal muscle atrophy typically first affects the face, shoulders, and arms, and then descends into the trunk and lower extremities.

As our understanding of FSHD pathophysiology has deepened, therapeutic efforts to prevent or delay FSHD disease progression have evolved from attempts to generally improve muscle function to approaches that specifically modulate DUX4 itself and the pathways that underlie DUX4 toxicity [12, 13]. With targeted therapies beginning to enter clinical trials [14, 15] it is essential to have robust tools with which to assess their effectiveness. Biomarkers are one such tool. Because many of the proposed disease-modifying therapies for FSHD selectively target DUX4, biomarkers designed to report on DUX4 level and/or activity are of critical importance in clinical development. Indeed, biomarker identification and validation has been a major goal of recent FSHD clinical trial workshops [16, 17]. To date, several studies have examined DUX4 target gene expression as a proxy for DUX4 activity from invasive muscle biopsies guided by MRI imaging [18–20]. However, circulating blood-based molecular biomarkers are of particular interest as they have the potential to provide rapid, objective, minimally invasive, and quantitative measurements that can be assayed repeatedly over time using typically inexpensive methods, in contrast to tissue biomarkers that require repeated muscle biopsies. A circulating biomarker may also allow physiological changes to be detected at a time when functional differences may not yet be measurable. Additionally, since all skeletal muscles are exposed to the circulation, muscle-derived serum or plasma proteins have the potential to reflect average disease burden throughout the body [21], “smoothing out” the local spatial variability in DUX4 expression observed with needle biopsies. However, since only a small fraction of muscles may present with active DUX4 expression at any one time in FSHD, a useful circulating biomarker may require a highly sensitive assay and little to no background contribution from healthy muscle and non-muscle tissue.

Unfortunately, there are limited data exploring peripheral blood biomarkers in FSHD and no independently validated circulating markers that can be used as predictive or prognostic tools. A study using whole transcriptome analysis to interrogate blood RNA expression profiles did not find any gene expression differences between individuals with FSHD and healthy controls that were significant after multiple hypothesis correction [22]. Studies using serum or plasma from FSHD patients and controls have identified proteins related to non-specific muscle damage [21, 23], immunity mediators [24–28], and miRNAs [26, 29, 30] as enriched in the disease context, but none were FSHD-specific and therefore might not reflect active DUX4-mediated disease processes. Although the proteomic panels used in prior studies in some cases incidentally included DUX4 targets, no studies to date have by design measured serum protein levels of DUX4 targets – despite the fact that quantifying DUX4 target expression in muscle is the only validated way to detect and track DUX4 activity [18, 19, 31]. While prior shotgun proteomics studies of FSHD serum [24, 26] could in principle have detected alterations in levels of DUX4 targets, more sensitive targeted assays may offer improved power to do so.

Here, we performed a focused and targeted study to determine whether any already well-established DUX4-regulated proteins would show differential levels in FSHD patient versus healthy control serum using a commercial Proximity Extension Assay proteomics technology from Olink Proteomics with the ultimate goal of identifying markers warranting further investigation. Most established DUX4 targets are not present on any of the Olink Proteomics biomarker panels, but because we are using DUX4 target expression levels as a proxy for DUX4 activity – rather than making suppositions that any particular target plays a direct role in FSHD pathophysiology – this need not be an impediment. Among the three DUX4 targets present on Olink Proteomics panels, we found that levels of placental alkaline phosphatase (ALPP) correlated with DUX4 expression in a cellular myoblast model of FSHD and in FSHD patient-derived myotube cell cultures. However, ALPP levels did not correspond to FSHD disease status or severity in human serum samples. Therefore, ALPP, as measured by this platform, does not appear to be a promising FSHD serum biomarker candidate but does have utility as a tool to evaluate DUX4 activity in discovery research.

## MATERIALS AND METHODS

### Cell culture

MB135 (Control-A), 54-1 (Control-B), MB073 (FSHD-A, FSHD type 1), 54-2 (FSHD-B, FSHD type 1), MB200 (FSHD-C, FSHD type 2), and MB135-iDUX4 immortalized human myoblasts were a gift from Dr. Stephen Tapscott and originated from the FSHD Research Center at the University of Rochester Medical Center. MB135-iDUX4 cells have been described previously [32]. All cell lines were authenticated by karyotype analysis and determined to be free of mycoplasma by PCR screening. Cell line characteristics are provided in Supplementary Table 1. Myoblasts were maintained in Ham’s F-10 Nutrient Mix (Gibco) supplemented with 20% Fetal Bovine Serum (Gibco), 10 ng/mL recombinant human basic fibroblast growth factor (Promega), and 1 μM dexamethasone (Sigma-Aldrich). MB135-iDUX4 myoblasts were additionally maintained in 2 μg/mL puromycin dihydrochloride (VWR). Induction of the DUX4 transgene was achieved by culturing cells in 1 μg/mL doxycycline hyclate (Sigma-Aldrich). Differentiation of myoblasts into myotubes was achieved by switching the fully confluent myoblast monolayer into Dulbecco’s Modified Eagle Medium (Gibco) containing 1% horse serum (Gibco) and Insulin-Transferrin-Selenium (Gibco). All cells were incubated at 37 °C with 5% CO_2_.

### Human serum

All serum samples (n = 20 healthy controls and n = 20 FSHD patients) were obtained following informed, written consent through the FSHD Research Center at the University of Rochester Medical Center under a local IRB-approved protocol and were deidentified. Donor characteristics are described in Supplementary Table 2.

### Preparation of cell lysates and supernatants for Olink Proteomics analysis

Myoblasts were seeded at a density of 2.5x10^5^ cells/well (MB135, 54-1, MB073, 54-2, MB200) or 1.5x10^5^ cells/well (MB135-iDUX4) on 12-well plates. Twenty-four hours prior to harvest, cells were washed three times with PBS and serum-free media was added. After 24 hours in serum-free media, supernatant and cell lysate were harvested as follows. The supernatant was removed, centrifuged for 5 minutes at 300 rcf to pellet any cell debris, transferred to a microcentrifuge tube, snap frozen with liquid nitrogen, and stored at -80 °C. Fifty microliters of ice-cold 1X RIPA Lysis Buffer (50 mM Tris-HCl pH 7.4, 150 mM NaCl, 1 mM EDTA, 1% Triton X-100, 0.1% sodium deoxycholate) supplemented with cOmplete Mini EDTA-free Protease Inhibitor Tablets (Roche) was added to each well, lysate was collected with a cell scraper, transferred to a microcentrifuge tube, incubated on ice for 15 minutes, sonicated with a Bioruptor (Diagenode) for 5 minutes on low (30 seconds on, 30 seconds off) to aid lysis, centrifuged for 5 minutes at 16,000 rcf at 4 °C, transferred to a new microcentrifuge tube, quantified using the BCA Protein Assay Kit (Pierce), snap frozen with liquid nitrogen, and stored at -80 °C. Samples were shipped on dry ice to Olink Proteomics.

### Olink Proteomics assay

Olink Proteomics conducted targeted, high-throughput, multiplex immunoassays of protein levels using the provided cell lysates, supernatants, and serum. Samples were run on the Target 96 Development (v.3511) and/or Target 96 Oncology III (v.4001) panels – these panels were selected because they were the only to include DUX4 targets. The assay readout of normalized protein expression (NPX) values [33] – log2-scaled scores with additional Inter-Plate Control normalization – was obtained from Olink Proteomics for downstream analysis. Because NPX scores are on a log2 scale, fold changes are computed as 2^(difference^ ^in^ ^NPX)^. The data from Olink Proteomics also included a limit of detection (LOD) score for each protein, defined to be the average score for negative control wells (buffer only) plus three standard deviations. Data below the LOD were used “as-is”, and although the response beneath the LOD can be non-linear, the effect of this on estimated fold changes is typically conservative (https://www.olink.com/faq/how-is-the-limit-of-detection-lod-estimated-and-handled).

### siRNA transfections

Silencer Select siRNAs were obtained from Thermo Fisher Scientific and transfected into MB135-iDUX4 myoblasts 36 hours prior to doxycycline induction using Lipofectamine RNAiMAX Transfection Reagent (Invitrogen) according to the manufacturer’s instructions. The siRNAs used in this study are listed below:

siCTRL: Silencer Select Negative Control No. 1 siRNA

siDUX4: CCUACACCUUCAGACUCUATT (sense), UAGAGUCUGAAGGUGUAGGCA (antisense)

### RNA isolation and RT-qPCR

Total RNA was extracted from whole cells using TRIzol Reagent (Invitrogen) following the manufacturer’s instructions. Isolated RNA was treated with DNase I (Invitrogen) and reverse transcribed into cDNA using SuperScript III Reverse Transcriptase (Invitrogen) and random hexamers (Invitrogen) according to the manufacturer’s protocol. Quantitative PCR was carried out on a CFX384 Touch Real-Time PCR Detection System (Bio-Rad) using primers specific to each gene of interest and iTaq Universal SYBR Green Supermix (Bio-Rad). The expression levels of target genes were normalized to that of the reference gene *RPL27* using the delta-Ct method [34]. The primers used in this study are listed below:

CKM F: CACCCCAAGTTCGAGGAGAT

CKM R: AGCGTTGGACACGTCAAATA

DUX4 transgene F: TAGGGGAAGAGGTAGACGGC

DUX4 transgene R: CGGTTCCGGGATTCCGATAG

KHDC1L F: CACCAATGGCAAAGCAGTGG

KHDC1L R: TCAGTCTCCGGTGTACGGTG

MYOG F: GCCAGACTATCCCCTTCCTC

MYOG R: GAGGCCGCGTTATGATAAAA

RPL27 F: GCAAGAAGAAGATCGCCAAG

RPL27 R: TCCAAGGGGATATCCACAGA

ZSCAN4 F: TGGAAATCAAGTGGCAAAAA

ZSCAN4 R: CTGCATGTGGACGTGGAC

### Statistical analysis

Data analysis and statistical tests were performed using GraphPad Prism software (version 9.0) or in the R programming environment using LIMMA [35] with plots generated using Prism or ggplot2 [36]. LIMMA computes empirical Bayes moderated t-statistics and F-statistics, with variances that are shrunken toward a global mean-variance trend (option trend = T); this helps stabilize results, particularly for studies in which the number of replicates is small. The focus of this study was on the small number of pre-defined DUX4 target proteins (from Table S1 of [31]) that were included on Olink Proteomics biomarker panels (one or two DUX4 target proteins on each of two 92-protein panels). In exploratory analyses of the full panels, false discovery rate (FDR) was used to control for multiple hypothesis testing.

## RESULTS

### Identification of DUX4 target genes on Olink Proteomics biomarker panels

We sought to identify candidate DUX4-induced genes for which commercial assays were readily available, with a particular interest in those with low background expression in somatic tissues (based on Genotype-Tissue Expression (GTEx) Project data, www.gtexportal.org) and those whose protein products were predicted to be secreted (based on Human Protein Atlas (HPA) secretome data [37], www.proteinatlas.org). We compared a set of 213 robust DUX4-upregulated target genes (Table S1 of [31]) to the 1,160 unique human protein targets found on 14 different Olink Proteomics Target 96 biomarker panels (**Figure 1A**). Three targets were present on both lists: placental alkaline phosphatase (ALPP), carbonic anhydrase 2 (CA2), and corticotropin releasing hormone binding protein (CRHBP). Of these, ALPP and CRHBP were included in the HPA secretome database and are predicted to encode signal peptides (as determined by UniProt annotation), making them excellent candidates to be secreted and have potential utility as FSHD blood-based molecular biomarkers.

**Figure 1.**
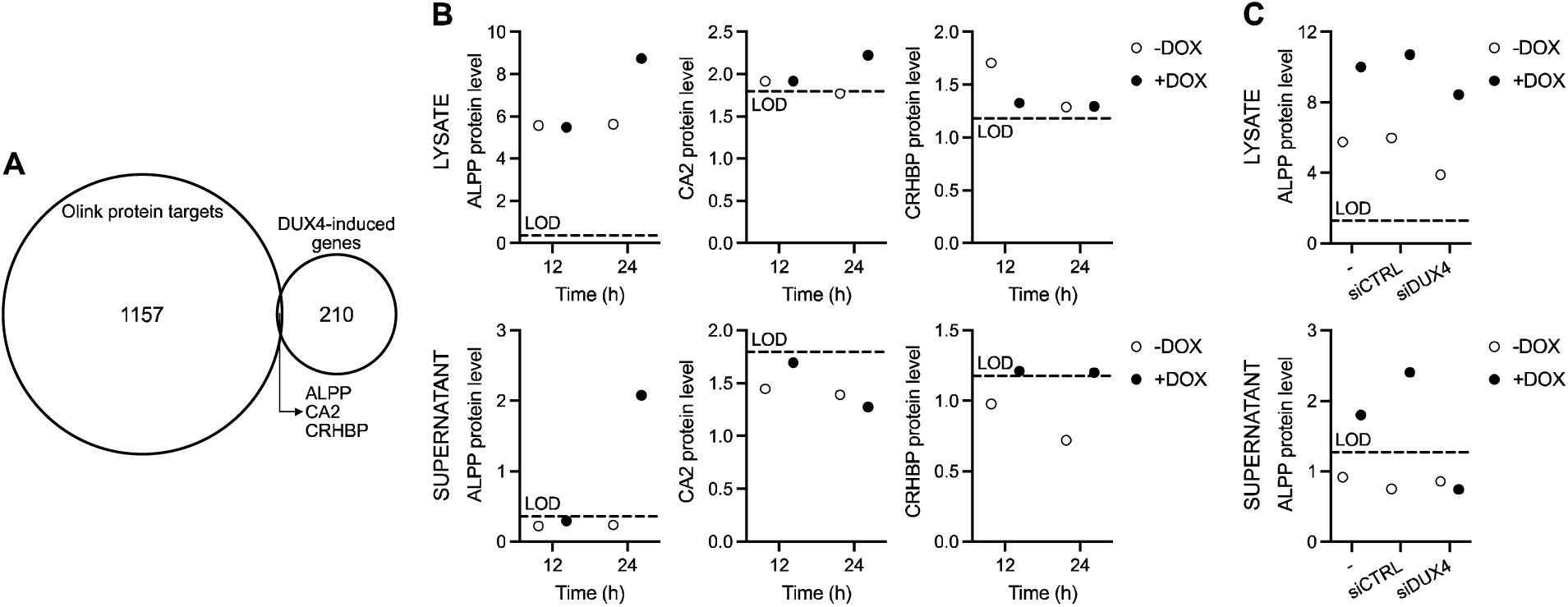
DUX4-induced ALPP protein is present in muscle cell supernatant. (**A**) Venn diagram showing the overlap between unique protein targets on the Olink Proteomics human Target 96 panels and DUX4-induced genes as classified in [31]. (**B**) The protein level of ALPP (right), CA2 (middle), and CRHBP (left) as measured by Olink Proteomics assay in cell lysate (top) and supernatant (bottom) harvested from MB135-iDUX4 myoblasts left untreated (-DOX) or treated with doxycycline (+DOX) to induce *DUX4* transgene expression for 12 or 24 hours. (**C**) ALPP protein levels as measured by Olink Proteomics assay in cell lysate (top) and supernatant (bottom) from MB135-iDUX4 myoblasts treated with or without doxycycline for 24 hours following transfection with no (-), non-targeting control (siCTRL), or DUX4 (siDUX4) siRNA. Protein levels in (**B**) and (**C**) are presented as normalized log2 expression values. LOD, limit of detection.

### ALPP distinguishes DUX4-expressing samples from controls in inducible DUX4 myoblasts

To measure the level of ALPP, CA2, and CRHBP in DUX4-expressing cells by Olink Proteomics assay we used the MB135-iDUX4 myoblast cell line, which has been engineered to inducibly express a *DUX4* transgene upon addition of doxycycline [32]. MB135-iDUX4 myoblasts were left untreated or cultured with doxycycline for 12 or 24 hours. Doxycycline treatment led to robust, time-dependent *DUX4* transgene induction as well as activation of DUX4 target genes [31] such as *KHDC1L* (**Supplementary Figure 1A**). Olink Proteomics analysis was performed on cell lysate and cell supernatant samples at both time points and normalized protein expression (NPX) values were reported on a log2 scale (**Figure 1B**, **Supplementary Figure 2A**). ALPP protein levels were above the limit of detection (LOD) in all cell lysate samples, with NPX values of ∼5.6 in both untreated and doxycycline-treated cells at 12 hours and also in untreated cells at 24 hours, but increasing to an NPX value of 8.7 in doxycycline-treated cells at 24 hours. Note that because NPX scores are on a log2 scale, the increase from ∼5.6 to 8.7 corresponds to a 2^3.1^ = 8.6-fold increase in protein level; other references to fold changes are based on similar calculations of 2^(delta^ ^NPX)^. In contrast, the level of CA2 and CRHPB protein in MB135-iDUX4 cell lysate hovered around the LOD, with CRHBP showing no correlation to DUX4 expression and CA2 displaying only a 1.4-fold increase after treatment with doxycycline for 24 hours. The inducible promoter in this cell system does exhibit some “leakiness” (**Supplementary Figure 1B**) [38], which may have contributed to the measurable cell lysate ALPP levels in the absence of doxycycline. In cell supernatants, CA2 and CRHBP protein levels were at or below the LOD, as were ALPP levels for samples left untreated or treated with doxycycline for 12 hours. However, after 24 hours of doxycycline treatment ALPP protein in cell supernatant was above the LOD and was 3.5-fold higher than the paired untreated sample. Because the NPX score for the untreated sample was below the LOD it may include a sizable contribution from non-specific background signal, so this fold change may be underestimated (see Methods); a similar caveat applies to other scores below the LOD. These results demonstrate that while CRHBP protein levels do not track with DUX4 expression, and CA2 levels only distinguish the highest DUX4-expressing condition from the rest in cell lysate, ALPP protein levels robustly correlate with DUX4 expression in both cell lysates and cell supernatants from MB135-iDUX4 myoblasts. Therefore, ALPP appeared to be a molecule worth pursuing further as a circulating FSHD biomarker.

To confirm that ALPP protein levels in MB135-iDUX4 cells are dependent on DUX4 expression, we knocked down the *DUX4* transgene using siRNA-mediated depletion and measured ALPP protein in cell lysate and supernatant in untreated cells or after 24 hours of doxycycline via Olink Proteomics assay (**Figure 1C**, **Supplementary Figure 2B**). Recapitulating our previous results, myoblasts treated with doxycycline to induce DUX4 for 24 hours showed elevated ALPP levels compared to untreated cells (∼20-fold in lysate, ∼3-fold in supernatant) when left untransfected or transfected with a non-targeting control siRNA. Also, as before, only in supernatant were ALPP levels in untreated cells below the LOD. This independent replication of prior ALPP measurements reveals low variability across replicates. Importantly, in doxycycline-treated cells transfected with siRNAs targeting the *DUX4* transgene, ALPP levels decreased (∼4-fold in lysate, ∼2.5-fold in supernatant) and in supernatant fell below the LOD. *DUX4* transgene and DUX4 target gene mRNA levels decreased >65% upon treatment with *DUX4* siRNAs, showing the efficacy of the knockdown (**Supplementary Figure 1C**). As a control to confirm that doxycycline treatment alone does not impact ALPP expression, we treated parental MB135 myoblasts with doxycycline and saw no effect on the level of ALPP, which hovered at or below the LOD, in cell lysate or supernatant (**Supplementary Figure 1D**). Together, these results demonstrate that ALPP protein detected in the supernatant of MB135-iDUX4 myoblasts via Olink Proteomics distinguishes DUX4-expressing samples from controls in an inducible DUX4 myoblast system.

### ALPP distinguishes FSHD from control myotubes

To determine if ALPP protein could distinguish cells expressing endogenous levels of DUX4 from controls, we employed two healthy and three FSHD patient-derived myoblast cell lines that were differentiated into myotubes for 2, 4, or 6 days (**Supplementary Figure 2C**). DUX4 levels are known to increase over this differentiation time course [39]. ALPP levels in cell lysate and supernatant were below the LOD in both control cell lines at most time points, whereas by day 2 (in lysate) or day 6 (in supernatant) of differentiation ALPP was above the LOD in all three FSHD cell lines (**Figure 2A**). In supernatant, this increase occurred gradually, with one of the FSHD cell lines showing measurable ALPP levels at day 2 of differentiation, two at day 4, and all three at day 6. Elevation of ALPP in FSHD versus control myotubes was significant (p < 0.05 by LIMMA moderated t-test) at all three time points for lysate (day 2: p = 0.038; day 4: p = 0.016; day 6: p = 0.0012) but just at day 6 for supernatant (day 2: p = 0.26; day 4: p = 0.084; day 6: p = 0.018). The myogenic genes *MYOG* and *CKM* were induced over this course of differentiation in all cell lines, as expected (**Supplementary Figure 3**), as was the DUX4 target gene *ZSCAN4* in the FSHD lines (**Figure 2B**). Note that *ZSCAN4* levels typically had the same rank order among FSHD cell lines (FSHD-C ≥ FSHD-B ≥ FSHD-A) as ALPP levels, consistent with both being induced by DUX4, but with just three samples such a concordance in ranks could also plausibly occur by chance. In conclusion, ALPP expression robustly distinguishes DUX4-expressing samples from controls in supernatants and lysates from differentiating muscle cell lines.

**Figure 2.**
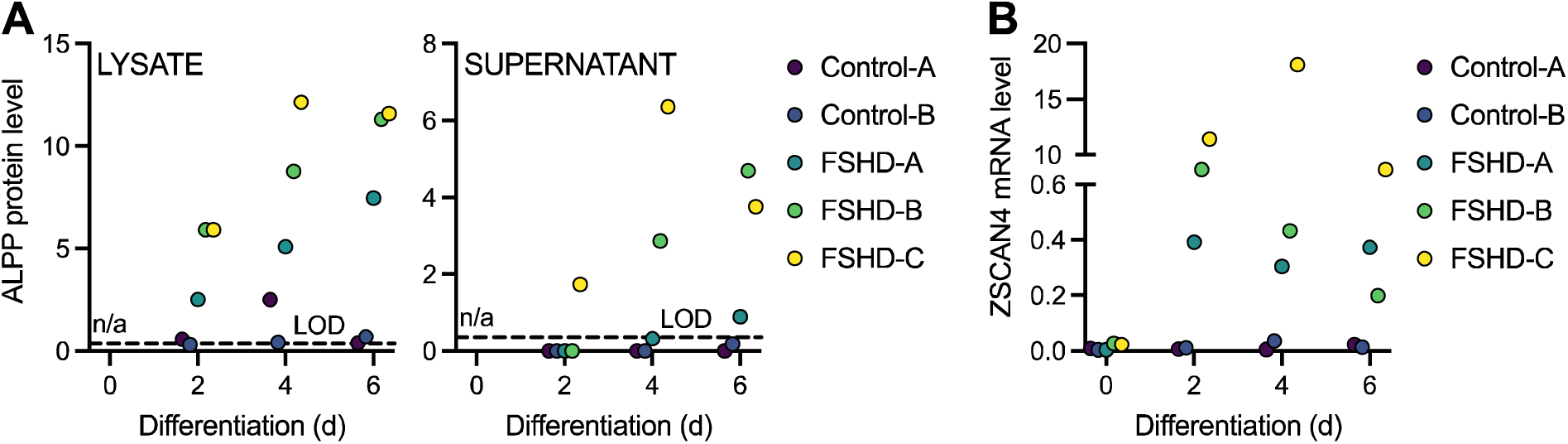
ALPP protein levels increase upon FSHD myoblast differentiation. (**A**) The normalized log2 expression level of ALPP protein as measured by Olink Proteomics assay in the cell lysate (left) and supernatant (right) of multiple control and FSHD myoblast cell lines differentiated into myotubes for 2, 4, or 6 days. (**B**) DUX4 target gene *ZSCAN4* mRNA levels as measured by RT-qPCR in control and FSHD myoblast cell lines differentiated into myotubes for 0, 2, 4, or 6 days. LOD, limit of detection. n/a, not available.

### ALPP levels do not distinguish FSHD from control serum

To measure the level of ALPP protein in blood from individuals with FSHD and healthy controls, we obtained 40 serum samples from the FSHD Research Center at the University of Rochester Medical Center and performed Olink Proteomics assays (**Supplementary Figure 4**). We fit a LIMMA linear model with factors for sex and disease status and their interaction (moderated two-way ANOVA) for the 92 proteins on the Target 96 Oncology III panel that includes ALPP. Here we summarize the results for ALPP, and results for all proteins are discussed in the Supplementary Material. There was not overall a significant association of ALPP protein levels in serum with FSHD status and/or sex (p-value = 0.104 for F-test) (**Figure 3A-B**), a result in contrast to our findings from the MB135-iDUX4 cellular myoblast model of FSHD and FSHD patient-derived myotube cell cultures. ALPP levels were on average higher in FSHD than control samples among the female subjects (increase of 1.14 NPX or ∼2.2-fold; nominal p-value = 0.018 for this contrast), but as the overall F-test was non-significant this should be regarded with caution (**Figure 3B**). There was also not a significant correlation between serum ALPP levels and age (rho = 0.29 and p-value = 0.07 by Spearman rank correlation test) (**Figure 3C**), or with disease severity among those with FSHD (rho = 0.01 and p-value = 0.97 by Spearman rank correlation test) (**Figure 3D**). The plot of FSHD clinical severity score (CSS) versus ALPP protein levels may have some hint of a V-shape but this could be a chance occurrence and is non-significant (p-value = 0.22 using a Hoeffding D-test for a not-necessarily monotonic association). Notably, serum ALPP levels were high at baseline even in healthy controls, suggesting a circulating, non-muscle source of ALPP protein. Indeed, ALPP is present in lung, gastrointestinal tract, cervix, and placenta [40], which may confound the circulating contribution from diseased muscle. Together, these results suggest that ALPP, as measured by the Olink Proteomics platform, is not a sensitive biomarker candidate for FSHD, though it could have utility as a tool in discovery research given its sensitivity and specificity in reporting out DUX4 activity in cell-based FSHD model systems.

**Figure 3.**
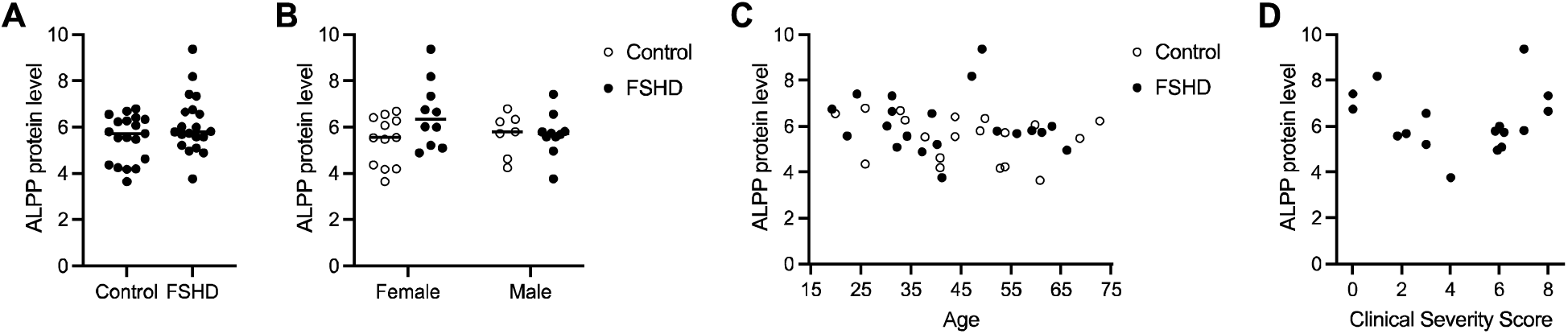

Serum ALPP levels do not predict FSHD disease state. (**A**) ALPP protein levels presented as normalized log2 expression values measured by Olink Proteomics assay in serum from 20 individuals with FSHD and 20 controls. (**B**-**D**) ALPP protein levels shown in (**A**) stratified by sex (**B**), age (**C**), and FSHD Clinical Severity Score (**D**).

## DISCUSSION

As potential FSHD therapies enter clinical trials, having quantitatively sensitive and rapidly responsive molecular biomarkers is critical for assessment of therapeutic approaches. Circulating biomarkers in particular provide the promise of reflecting overall disease burden in ways not possible using physical examination, tissue biopsy, or imaging methods. In this study, we used a commercial platform provided by Olink Proteomics to measure protein levels of three known DUX4-induced targets – ALPP, CA2, and CRHBP – in lysate and supernatant from a cellular myoblast model of FSHD and in FSHD patient-derived myotube cell cultures. The two cell-based systems revealed ALPP as a promising secreted FSHD biomarker candidate, so we performed Olink Proteomics validation assays for ALPP in serum from individuals with FSHD and healthy controls, but these did not show a correlation between serum ALPP levels and disease state or severity and revealed high background levels of ALPP in human serum. Therefore, we conclude that ALPP, as detected by the Olink Proteomics platform, is not a good clinical FSHD biomarker despite its promising performance in cellular systems.

Our initial examination of background tissue expression was primarily based on RNA expression levels from GTEx, which may not reflect serum protein levels. Moreover, the GTEx database does not include placenta, an organ where ALPP is expressed based on data from the HPA. Other confounding systemic non-muscle sources of ALPP may arise from the lung, gastrointestinal cells, and/or the cervix [40]. However, the Human Plasma Proteome Project (HPPP) database (www.hupo.org/plasma-proteome-project) showed little evidence for plasma ALPP expression at the time of this study, and the HPA page for ALPP says that it is not detected in plasma by mass spectrometry based on the PeptideAtlas database (build id=465). We note however that HPA now also includes Proximity Extension Assay data from plasma based on a recent study using Olink Proteomics [41], and this does show fairly high levels of ALPP among healthy controls, consistent with our observations. It may be possible that ALPP homologs could be contributing to the observed background serum ALPP levels, as we were unable to assess the specificity of the Olink Proteomics assay for ALPP (versus, for example, ALPG) and therefore cannot dismiss the possibility that the assay may be cross-reacting with other proteins.

The prolonged, non-linear muscle degradation and highly variable clinical presentation typical of FSHD requires precise measurement of biomarker concentrations so that subtle changes in disease can be detected. The Olink Proteomics technology used in this study claims high specificity and sensitivity and could provide a powerful tool for quantifying low-abundance DUX4-induced proteins that might have been otherwise thought too variable to be useful biomarkers for disease assessment. In this study we were limited to a small number of DUX4-induced proteins that were present on existing Olink Proteomics panels. There is the possibility of designing custom Olink Proteomics panels that consolidate DUX4 target proteins and other FSHD-relevant proteins from existing panels onto smaller focused panels, but targeted mass spectrometry approaches using Parallel or Multiple Reaction Monitoring (PRM/MRM) offer an interesting alternative for profiling a larger collection of DUX4-induced proteins in serum, as has been done for microdialysates from FSHD muscle [24].

As our goal was to identify DUX4-induced serum biomarkers, we used only the two Olink Proteomics panels that included protein products of known DUX4-induced genes. The two panels used in the cell culture studies contained 181 other protein targets, and although not of primary interest we also examined their levels. Several increased in expression following doxycycline induction of DUX4 in the MB135-iDUX4 myoblasts (**Supplementary Figure 2**, Supplementary Table 3). Of these, *VMO1*, *PSPN*, *PTP4A1*, and *NOV* are upregulated at the mRNA level in DUX4-expressing cells [32], but were not present on our original list of 213 robust DUX4-upregulated target genes. Only PTP4A1 protein levels correlated with DUX4 expression in both MB135-iDUX4 cell lysates and cell supernatants; however, PTP4A1 did not distinguish FSHD from control myotubes, illustrating the importance of validating any findings derived from inducible DUX4 cell models with FSHD patient-derived muscle cells expressing endogenous levels of DUX4.

In the serum studies we used only one Olink Proteomics biomarker panel – the Target 96 Oncology III panel containing ALPP – since the two DUX4 targets on the Target 96 Development panel did not show clear DUX4-dependent induction in cell culture studies, which may be due in part to their levels being near or below the Olink Proteomics assay limit of detection. Incidental exploratory findings for the other proteins on the Target 96 Oncology III panel in serum are discussed in the legend to Supplementary Figure 4.

The decision to focus our circulating biomarker search on DUX4-induced genes was intended to ensure that any hits were clearly relevant to FSHD, but also required that the DUX4 target be secreted from muscle cells. Our time course differentiation data suggest that ALPP secretion requires high, sustained DUX4 (and DUX4 target) expression, a prerequisite that may eliminate other candidate molecules not detectable under the conditions used here. However, our study also clearly demonstrated the utility of ALPP as a discovery tool. Measuring ALPP levels in the supernatant of DUX4-expressing cells would allow for time course experiments not possible with currently used RNA and protein analysis methods.

Overall, it is possible that multiple assessment approaches will be necessary to evaluate changes in FSHD disease burden over the course of a clinical trial. Combining information from serum biomarkers, muscle biopsy, magnetic resonance imaging, and clinical strength and activity measurements may increase our ability to assess disease progression and evaluate FSHD therapeutics.

## Data Availability

All data produced in the present study are available upon reasonable request to the authors.

## ACKNOWLEDGEMENTS

We thank the FSHD families whose participation is critical for progress. This study was supported by the FSHD Society (S.J. and A.E.C.), the Geraldi Norton Foundation (S.J. and A.E.C.), NIH 5P50HD060848-15 Wellstone Center for FSHD (O.D.K.), and the RNA Bioscience Initiative at the University of Colorado Anschutz Medical Campus (S.J.). The Genotype-Tissue Expression (GTEx) Project was supported by the Common Fund of the Office of the Director of the National Institutes of Health and by NCI, NHGRI, NHLBI, NIDA, NIMH, and NINDS.

## AUTHOR CONTRIBUTIONS

A.E.C., J.A., O.D.K., and S.J. conceived and designed the study. A.E.C. performed experiments. R.T. provided critical reagents. A.E.C., J.A., O.D.K., and S.J. analyzed data. A.E.C. wrote the manuscript with input from all authors.

## SUPPLEMENTARY FIGURE LEGENDS

**Supplementary Figure 1.**
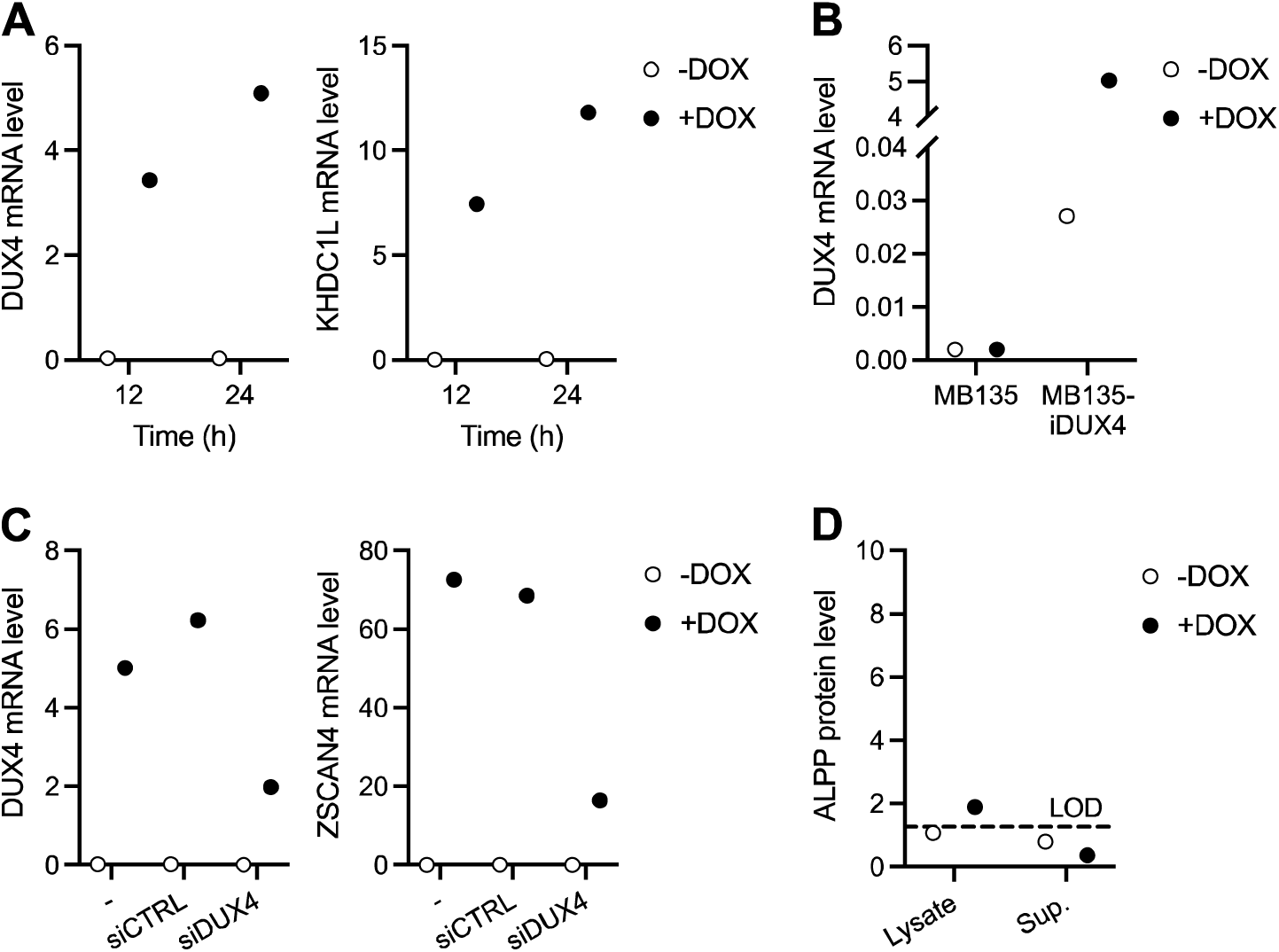
MB135-iDUX4 myoblasts as a model system to uncover secreted DUX4-induced proteins. (**A**) Level of *DUX4* and DUX4 target gene *KHDC1L* mRNA as measured by RT-qPCR in MB135-iDUX4 myoblasts left untreated (-DOX) or treated with doxycycline (+DOX) to induce *DUX4* transgene expression for 12 or 24 hours. (**B**) *DUX4* mRNA as measured by RT-qPCR in parental MB135 myoblasts and in MB135-iDUX4 myoblasts left untreated (-DOX) or treated with doxycycline (+DOX) for 24 hours. (**C**) *DUX4* and DUX4 target gene *ZSCAN4* mRNA levels measured by RT-qPCR in MB135-iDUX4 myoblasts treated with or without doxycycline for 24 hours following transfection with no (-), non-targeting control (siCTRL), or *DUX4* (siDUX4) siRNA. (**D**) ALPP protein levels (log2 normalized) as measured by Olink Proteomics assay in the cell lysate and supernatant (Sup.) of parental MB135 myoblasts treated with (+DOX) or without (-DOX) doxycycline for 24 hours.

**Supplementary Figure 2.**
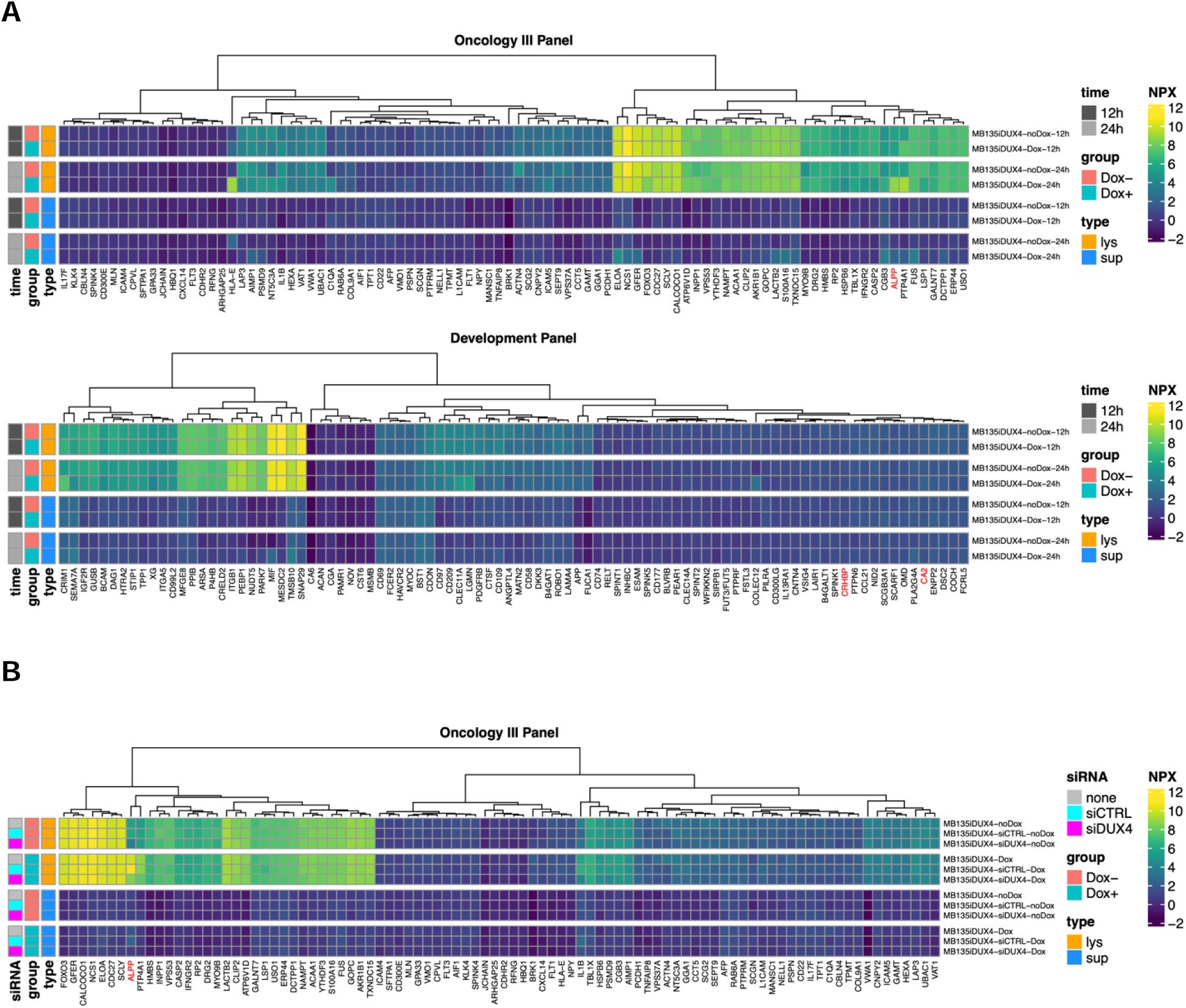
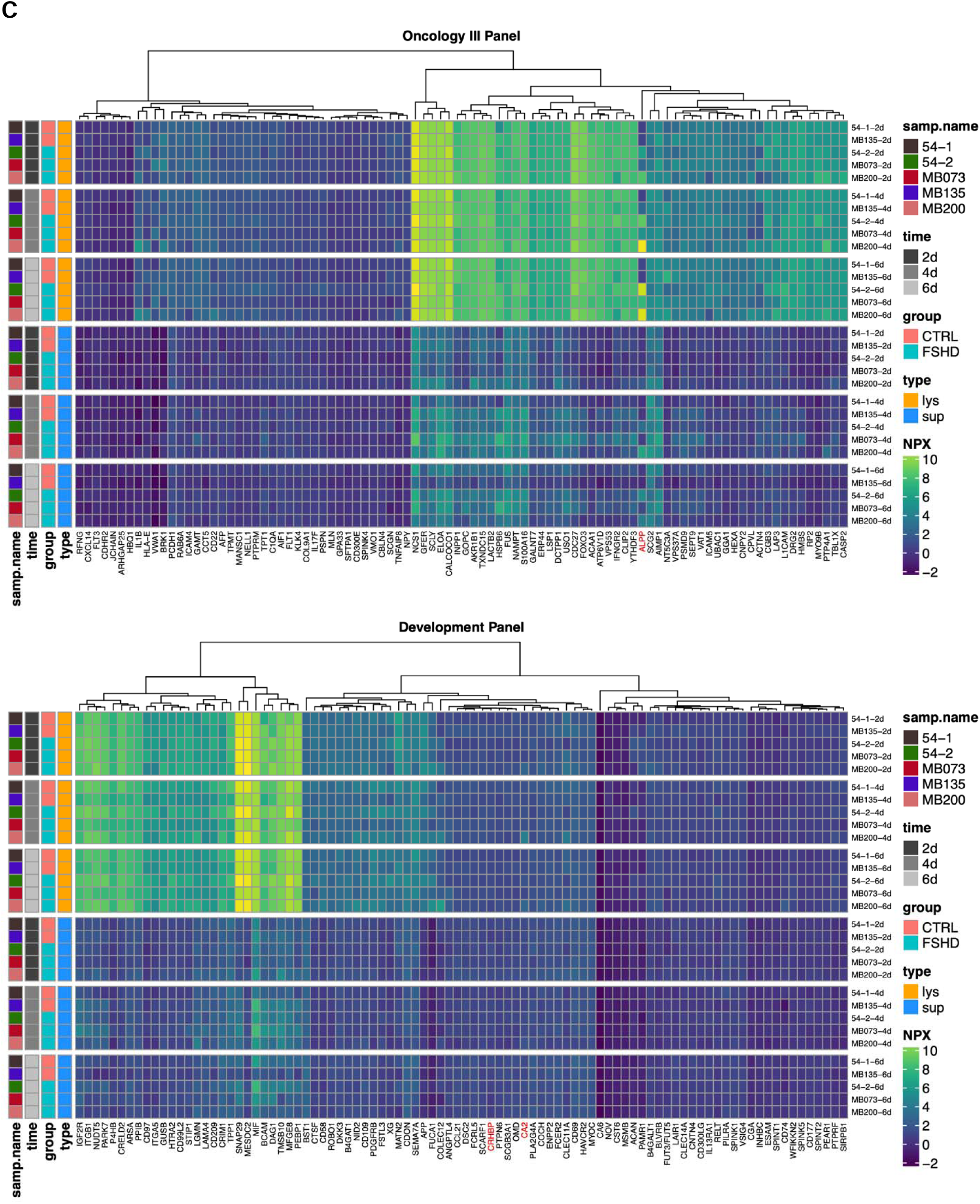
Protein levels from cell culture studies. Separate heatmaps are shown for the Olink Proteomics Target 96 Oncology III and Target 96 Development panels for the (**A**) MB135-iDUX4 data, (**B**) MB135-iDUX4 siRNA data, and (**C**) FSHD versus control myotube data. NPX scores are on a log2 scale. Sample type is indicated by columns in the left margin of each heatmap, with legends to the right of each heatmap. The DUX4 target proteins ALPP, CA2, and CRHBP are labelled in red in the lower margin. Dox-, untreated; Dox+, doxycycline-treated; lys, cell lysate; sup, cell supernatant.

**Supplementary Figure 3.**
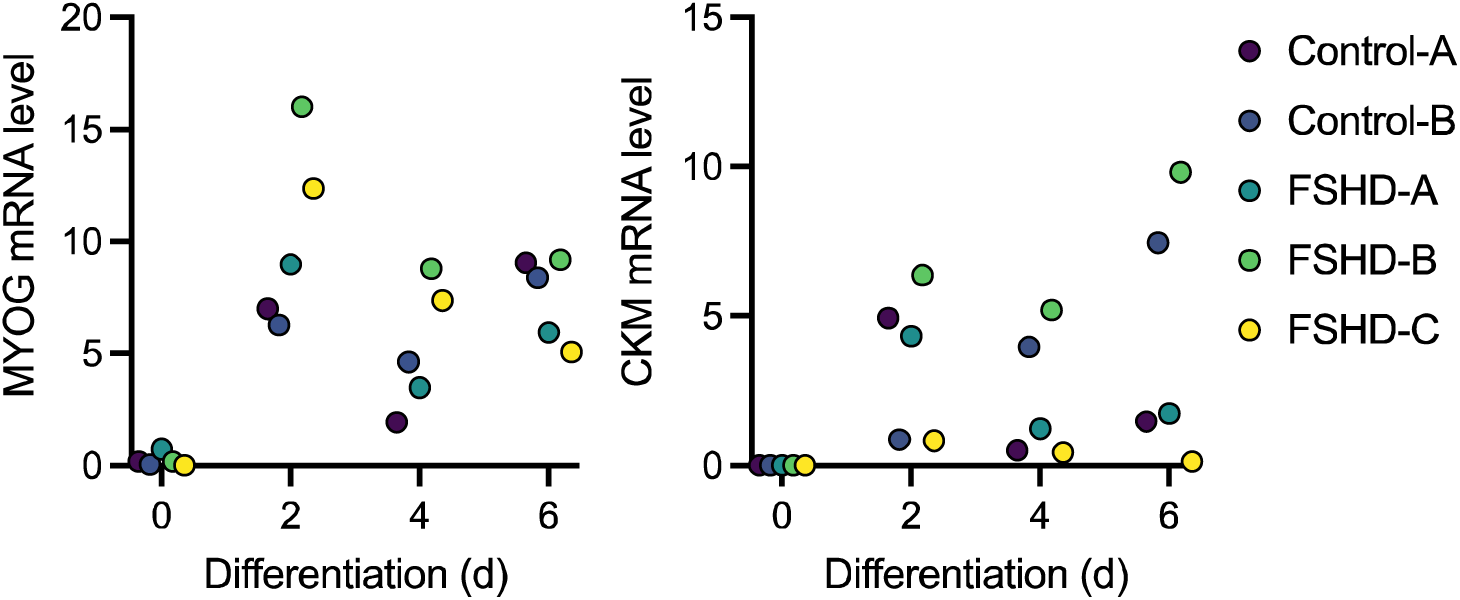
Control and FSHD cell lines differentiate into myotubes. mRNA level of myogenic genes *MYOG* and *CKM* as measured by RT-qPCR in multiple control and FSHD myoblast cell lines differentiated into myotubes for 0, 2, 4, or 6 days.

**Supplementary Figure 4.**
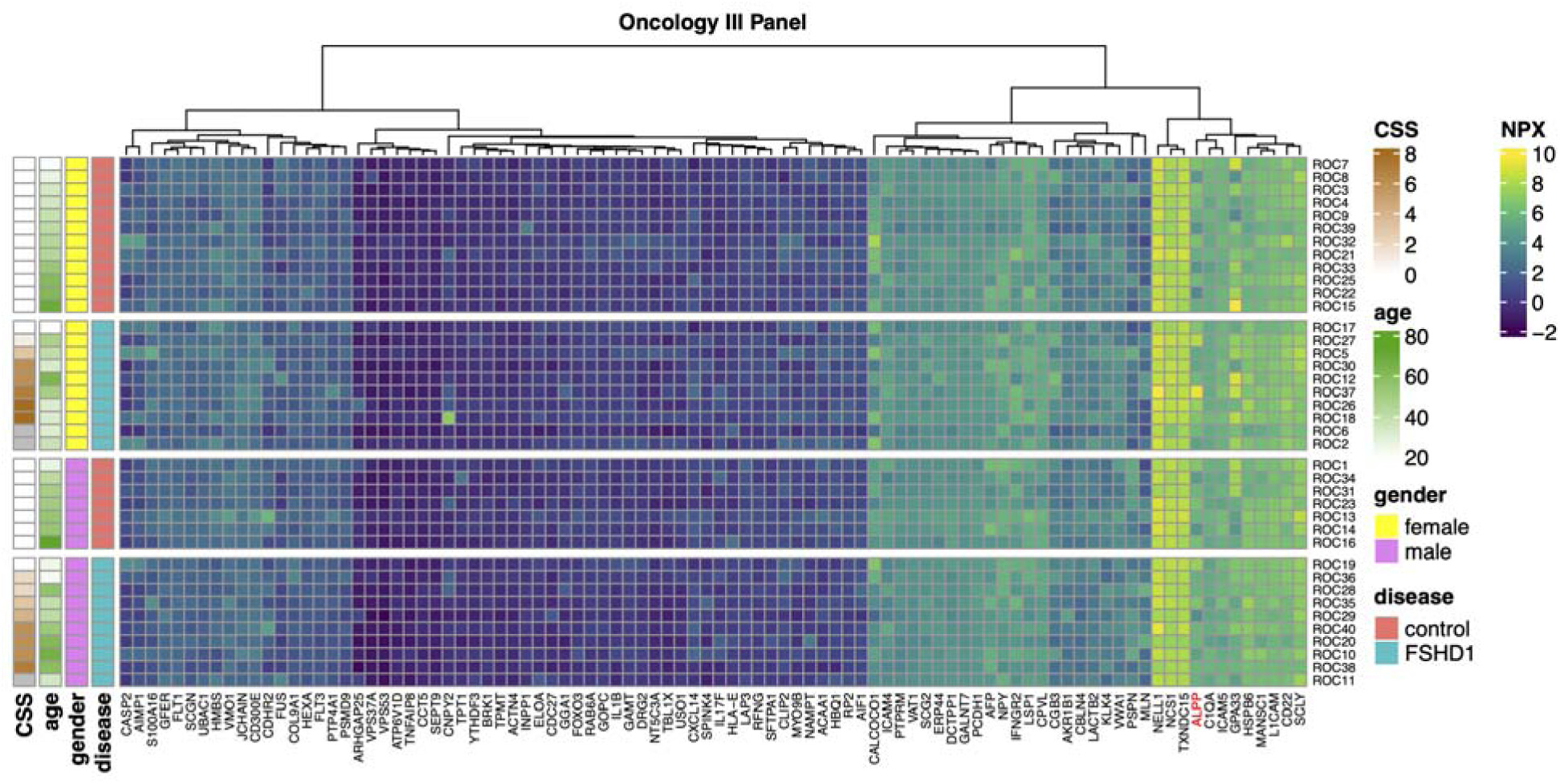
Protein levels from serum study. Only the Olink Proteomics Target 96 Oncology III panel was used for serum analysis. NPX scores are on a log2 scale. Sample metadata (disease status, sex, age, CSS) is indicated by columns in the left margin of the heatmap, with legends to the right of the heatmap. (Missing values of CSS are shaded grey.) The DUX4 target protein ALPP is labelled in red in the lower margin, and is discussed in detail in the main text. The only proteins showing a significant difference associated with disease status and/or sex at FDR < 0.1, based on a LIMMA moderated F-test that is analogous to a two-way ANOVA, were HSPB6 and PSPN, both with FDR = 0.034. HSPB6 was up ∼1.9-fold in FSHD versus control females with FDR = 0.021, and was up ∼1.3-fold in FSHD versus control males but with non-significant FDR = 0.63. PSPN was up ∼2.8-fold in males versus females with FDR = 0.0045, but without a significant interaction with FSHD status (FDR = 0.86). HSPB6, or Heat Shock Protein Family B (Small) Member 6, has recently been reported to be upregulated at the mRNA level in a transcriptomic study of Ant1-overexpressing mice [42], and overexpression of human ANT1 – a 4q35 gene involved in mitochondrial function – has also been associated with FSHD [43]. PSPN, or Persephin, is a secreted ligand of GDNF and TGF-beta proteins that is encoded on chromosome 19 and was also strongly elevated in male versus female plasma in a recent study using the Olink Proteomics platform [41].

## SUPPLEMENTARY TABLE

Supplementary Table 1. Myoblast cell line characteristics.

Supplementary Table 2. Serum donor characteristics.

Supplementary Table 3. Olink Proteomics normalized protein expression (NPX) values.

